# Documented clinical genetic testing among carriers of hereditary breast and ovarian cancer variants: Ancestry and socioeconomic disparities in the All of Us research program

**DOI:** 10.64898/2026.06.09.26355262

**Authors:** Srinivasulu Yerukala Sathipati, Hebbring Scott

**Affiliations:** Center for Precision Medicine Research, Marshfield Clinic Research Institute, Sanford Health-Marshfield Clinic, Marshfield, WI, USA

## Abstract

**Importance:** Hereditary breast and ovarian cancer (HBOC) variant carriers benefit from risk-reducing interventions, but only if identified. The extent to which carriers are clinically recognized, and whether recognition is equitable across diverse populations, is poorly characterized in a single large U.S. cohort.

**Objective:** To estimate P/LP HBOC carrier prevalence across genetic ancestry groups, quantify documented clinical genetic testing among carriers, and evaluate ancestry and socioeconomic disparities in testing.

**Design, Setting, and Participants:** Cross-sectional analysis of the All of Us Research Program Controlled Tier (Curated Data Repository v8/C2024Q3R9), comprising participants with short-read whole-genome sequencing and linked electronic health record (EHR) and survey data. Carriers were ascertained from research genomic data independent of clinical testing.

**Exposures:** Genetically inferred ancestry (African [AFR], Admixed American [AMR], East Asian [EAS], European [EUR], Middle Eastern [MID], South Asian [SAS]); self-reported household income and educational attainment.

**Main Outcomes and Measures:** (1) Carrier prevalence with Wilson 95% CIs; (2) documented clinical genetic testing (procedure codes) among carriers; (3) adjusted odds of documented testing among women, by ancestry, before and after socioeconomic adjustment, using multivariable logistic regression.

**Results:** Among 414,830 participants, P/LP HBOC carrier prevalence was 1.42% (95% CI, 1.38–1.45) overall and similar across ancestry groups (AFR 1.24%, AMR 1.32%, EAS 1.19%, EUR 1.52%, MID 1.68%, SAS 1.33%; overlapping CIs). Among 250,071 women in the testing analysis, documented clinical genetic testing was rare: only 74 of 5,878 carriers overall (1.3%) and 59 of 3,572 European-ancestry carriers (1.7%) had a documented test, with counts below reportable thresholds in all other ancestry groups. African-ancestry women had lower adjusted odds of documented testing than European-ancestry women (Model 1 adjusted odds ratio [aOR], 0.32; 95% CI, 0.27–0.39), an association that attenuated but persisted after adjustment for income and education (Model 2 aOR, 0.48; 95% CI, 0.40–0.58; P < 0.001); Admixed American women also had reduced adjusted odds (aOR, 0.71; 95% CI, 0.61–0.84). Lower income and lower education were independently and dose-dependently associated with lower testing odds (income <$25,000 aOR, 0.46; high-school education aOR, 0.54).

**Conclusions and Relevance:** High-risk HBOC variant carriers are present across all ancestry groups at similar frequencies, yet documented clinical genetic testing was disparate in the different ancestry groups. African-ancestry women experience a testing gap that is not fully explained by socioeconomic position, implicating structural barriers in access and referral. Population-level strategies that decouple carrier identification from current referral pathways may be required to close this gap.

**Key Points:** *Question:* Among carriers of pathogenic or likely-pathogenic hereditary breast and ovarian cancer (HBOC) variants identified from research whole-genome sequencing, how often is clinical genetic testing documented, and does it differ by genetic ancestry and socioeconomic position?

*Findings:* In this cross-sectional study of 414,830 All of Us participants with whole-genome sequencing, P/LP HBOC carrier prevalence was similar across genetic ancestry groups (∼1.4%), yet documented clinical genetic testing among carriers was rare (1.3% overall; 1.7% among European-ancestry carriers; suppressed in other groups). African-ancestry women had markedly lower adjusted odds of documented testing, a gap that persisted after adjustment for income and education.

*Meaning:* High-risk variant carriers are distributed across all ancestry groups, but the overwhelming majority are never clinically identified, and African-ancestry women face an access gap not explained by socioeconomic position alone.

## Background

Pathogenic and likely-pathogenic (P/LP) germline variants in hereditary breast and ovarian cancer (HBOC) genes—including BRCA1, BRCA2, PALB2, CHEK2, and ATM—substantially increase lifetime risk of breast, ovarian, and other cancers[1-4]. Identification of carriers enables evidence-based risk reduction, including enhanced surveillance, risk-reducing surgery, and cascade testing of relatives[3 5]. The clinical value of carrier status, however, is realized only when carriers are recognized.

The magnitude of this risk is substantial. Female BRCA1 and BRCA2 carriers face cumulative breast cancer risks of approximately 72% and 69% by age 80 years, and ovarian cancer risks of approximately 44% and 17%, respectively [6]; moderate-penetrance genes such as PALB2, ATM, and CHEK2 confer roughly 2-to 4-fold increases in breast cancer risk [1 7]. These variants are not rare cumulatively— pathogenic or likely-pathogenic HBOC variants are carried by approximately 1–2% of the general population [8 9] and account for a substantial share of breast and ovarian cancers [2 3]. Identification is, moreover, actionable: risk-reducing salpingo-oophorectomy lowers ovarian cancer risk by approximately 80% and is associated with reduced all-cause mortality, and risk-reducing mastectomy lowers breast cancer risk by approximately 90% [10]. The difference between a recognized and an unrecognized carrier can therefore be the difference between prevention and a preventable cancer.

Current ascertainment relies on phenotype-and family-history–based referral to clinical genetic testing[11]. Multiple studies indicate that this pathway misses the majority of carriers in the population[8 12] and does so unequally: Black and Hispanic individuals are consistently less likely to undergo germline testing than White individuals[13-15], reflecting differences in awareness, provider referral, geographic and language access, and socioeconomic resources[14]. Whether the shortfall is driven primarily by socioeconomic position or by factors that persist beyond it has direct implications for intervention design.

The All of Us Research Program[16] offers a unique opportunity to address these questions, pairing whole-genome sequencing with electronic health record and survey data in a large, ancestrally diverse U.S. cohort[16 17]. Because carriers can be ascertained directly from research genomic data—independent of whether they were ever clinically tested—the program permits direct measurement of the gap between who carries a high-risk variant and who has been clinically identified.

We therefore (1) estimated P/LP HBOC carrier prevalence across genetic ancestry groups, (2) quantified documented clinical genetic testing among carriers, and (3) evaluated whether disparities in documented testing by ancestry are explained by socioeconomic position.

## Methods

### Study Design and Population

We performed a cross-sectional analysis using the All of Us Research Program Controlled Tier (Curated Data Repository v8; C2024Q3R9)[16 17]. The analytic population comprised participants with short-read whole-genome sequencing (WGS) and a program-assigned genetic ancestry prediction. Analyses of documented genetic testing were restricted to participants with female sex at birth, consistent with the clinical context of HBOC testing. The study used de-identified data and was conducted under All of Us data-use policies; small-cell suppression (counts <20) was applied to all reported cells in accordance with program disclosure rules. The study is reported in accordance with the Strengthening the Reporting of Observational Studies in Epidemiology (STROBE) guideline for cross-sectional studies[18].

### Genomic Identification of Variant Carriers

Carriers were identified from the All of Us Cohort-Builder genomic tables in BigQuery. We evaluated an established HBOC gene panel (BRCA1, BRCA2, PALB2, CHEK2, ATM, TP53, PTEN, CDH1, STK11, RAD51C, RAD51D, BARD1) consistent with current clinical guidelines[3]. Qualifying variants were rare (allele frequency <1%)[9] and classified using ClinVar clinical-significance assertions[19] interpreted in line with American College of Medical Genetics and Genomics/Association for Molecular Pathology variant-classification standards[20]. Under the primary (strict) definition, a variant qualified only if its entire ClinVar label set was pathogenic and/or likely-pathogenic (allowing the non-informative “not provided”) with no benign, likely-benign, or uncertain-significance conflict. A sensitivity definition additionally rescued unambiguous loss-of-function variants (frameshift, stop-gain, canonical splice, start/stop-loss) that carried a P/LP assertion without benign/VUS conflict, recovering established founder alleles. Carrier status was defined as carriage of ≥1 alternate allele; zygosity was not distinguished. The distribution of carriers and qualifying variants across individual HBOC genes is provided in **eTable 1** in the Supplement. The panel was pre-specified as the NCCN HBOC genes because each is individually actionable for breast/ovarian cancer. In a sensitivity analysis we repeated carrier ascertainment using the cancer-phenotype subset of the ACMG Secondary Findings v3.2 list[21],both including and excluding MUTYH; because MUTYH-associated polyposis is autosomal recessive and only biallelic carriers are actionable, monoallelic MUTYH carriers — which predominate and are not distinguishable by zygosity here — were excluded in the primary comparison (**eTable 3**).

### Outcome and Phenotype Definitions

The primary outcome was documented clinical genetic testing, defined from EHR procedure codes for germline genetic testing and counseling. Breast and ovarian cancer diagnoses were defined from condition concepts using the OMOP concept hierarchy. Demographic variables were obtained from program data; household income and educational attainment were derived from the All of Us survey (‘The Basics’). Health-insurance survey items were not used because the survey instrument conflated multiple questions and yielded unreliable coding. Income was Unknown (the survey ‘Unknown’ category, pooling skip and prefer-not-to-answer responses) for 50,769 women (20.3%) and education attainment for 4,978 (2.0%); missing values were retained as an explicit category rather than excluded, so no participants were dropped for missing socioeconomic data.

### Covariates and Genetic Ancestry

Genetic ancestry was taken from program-provided ancestry predictions (AFR, AMR, EAS, EUR, MID, SAS), with European ancestry as the reference group. Models adjusted for age, any breast/ovarian cancer diagnosis, and carrier status. Genetic principal components were considered but excluded from the testing-disparity models: because the categorical ancestry label is derived from the same principal-component space, including both induced severe collinearity and unstable estimates, and adjusting a health-care– access outcome for fine-scale genetic structure constitutes over-adjustment of the exposure of interest. Principal components were therefore reserved for population-structure (prevalence) considerations only.

### Statistical Analysis

Carrier prevalence was estimated overall and by ancestry with Wilson 95% confidence intervals. Documented testing among carriers was tabulated by ancestry with small-cell suppression. Disparities in documented testing were evaluated among women using multivariable logistic regression in a prespecified nested sequence: Model 1 adjusted for ancestry, age, cancer diagnosis, and carrier status; Model 2 added household income and education. Attenuation from Model 1 to Model 2 quantified the degree to which ancestry differences were explained by socioeconomic position. A two-sided P<.05 defined statistical significance. Analyses used Python (statsmodels).

## Results

### Cohort Characteristics

The analytic cohort comprised 414, 830 participants with WGS and an assigned genetic ancestry (Table 1). European ancestry was most common (234, 353; 56.5%), followed by African (84, 148; 20.3%) and Admixed American (79, 106; 19.1%) ancestry, with smaller East Asian (10, 099), South Asian (5,579), and Middle Eastern (1,545) groups. The testing-disparity analysis included 250, 071 women with complete covariate data. Participant flow through the study is shown in **eFigure 1** in the Supplement.

**Table 1.** Characteristics of the study population, All of Us Research Program (v8).

| Characteristic | Value |
| --- | --- |
| Participants with WGS and genetic ancestry, No. | 414, 830 |
| Women in testing analysis, No. | 250, 071 |
| Age, mean (SD), years | 56.2 (17.0) |
| European ancestry, No. (%) | 234,353 (56.5) |
| African ancestry, No. (%) | 84,148 (20.3) |
| Admixed American ancestry, No. (%) | 79,106 (19.1) |
| East Asian ancestry, No. (%) | 10,099 (2.4) |
| South Asian ancestry, No. (%) | 5,579 (1.3) |
| Middle Eastern ancestry, No. (%) | 1,545 (0.4) |
| P/LP HBOC carriers (strict definition), No. (%) | 5,878 (1.42) |

*WGS indicates whole-genome sequencing; P/LP, pathogenic or likely-pathogenic;*
*HBOC, hereditary breast and ovarian cancer. Percentages are of the WGS cohort.*

### Carrier Prevalence Is Similar Across Genetic Ancestry Groups

Under the strict definition, 5878 carriers were identified (overall prevalence, 1.42%; 95% CI, 1.38–1.45). Prevalence was similar across ancestry groups, with overlapping confidence intervals (**Table 2**; **Figure 1**): African 1.24% (95% CI, 1.17–1.32), Admixed American 1.32% (1.24–1.40), East Asian 1.19% (1.00–1.42), European 1.52% (1.48– 1.58), Middle Eastern 1.68% (1.15–2.45), and South Asian 1.33% (1.06–1.66). High-risk carriers were thus distributed across all populations at broadly comparable frequencies.

**Table 2.** P/LP HBOC carrier prevalence by genetic ancestry (strict variant definition).

| **Genetic ancestry** | **N (WGS)** | **Carriers, No.** | **Prevalence, % (95% CI)** |
| --- | --- | --- | --- |
| Overall | 414,830 | 5,878 | 1.42 (1.38–1.45) |
| African | 84,148 | 1,044 | 1.24 (1.17–1.32) |
| Admixed American | 79,106 | 1,042 | 1.32 (1.24–1.40) |
| East Asian | 10,099 | 120 | 1.19 (1.00–1.42) |
| European | 234,353 | 3572 | 1.52 (1.48–1.58) |
| Middle Eastern | 1,545 | 26 | 1.68 (1.15–2.45) |
| South Asian | 5,579 | 74 | 1.33 (1.06–1.66) |

*Wilson 95% confidence intervals. Estimates use the strict P/LP variant definition; a loss-of-function–rescue sensitivity definition yielded consistent results.*

**Figure 1.**
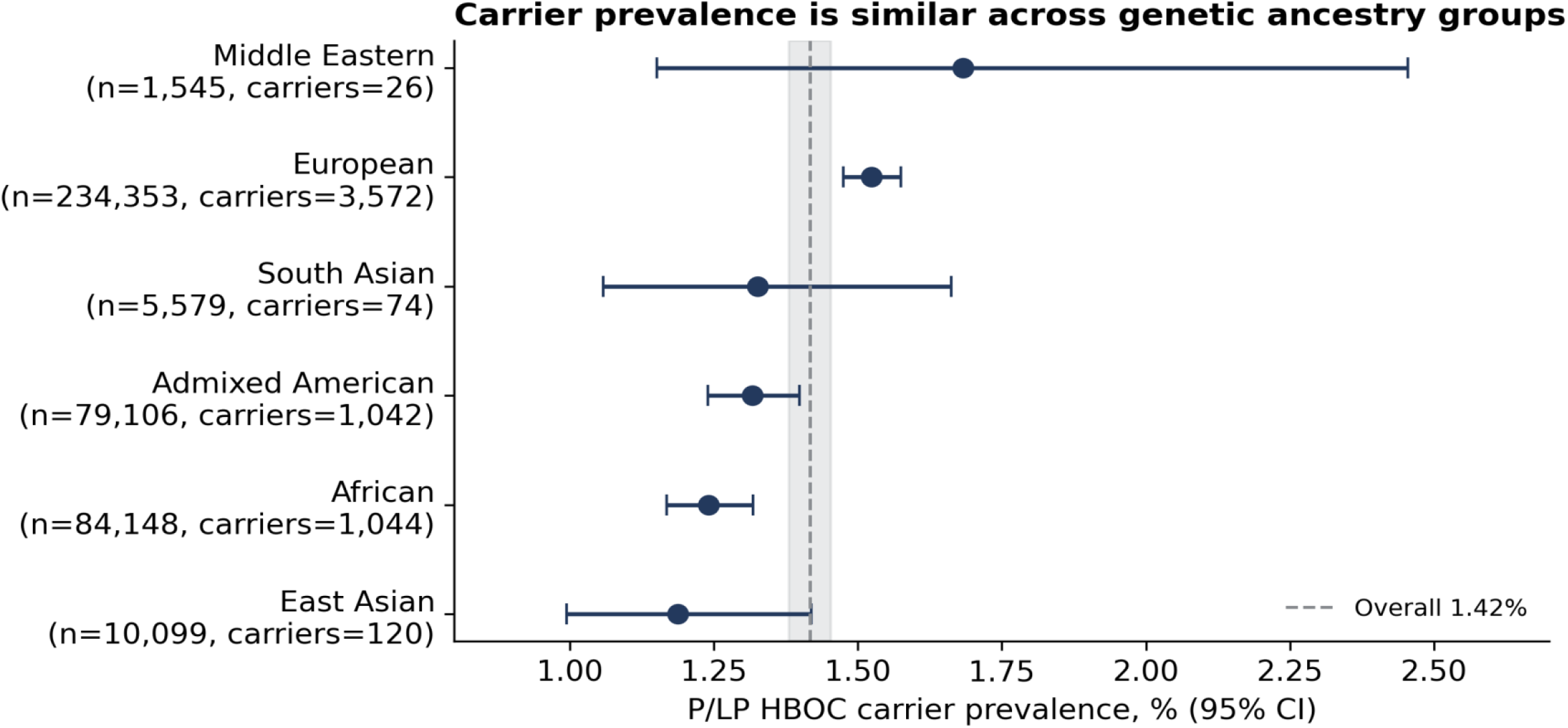
Carrier prevalence (%) with 95% confidence intervals by genetic ancestry. The dashed line and shaded band denote the overall estimate. Confidence intervals overlap across groups.

### Documented Clinical Genetic Testing Among Carriers Is Rare

Despite the substantial carrier burden, documented clinical genetic testing among known carriers was uncommon. Overall, only 74 of 5,878 carriers (1.3%) had a documented genetic test, including 59 of 3,572 European-ancestry carriers (1.7%); in every other ancestry group the number of tested carriers fell below the reportable threshold (<20) and was suppressed. A conservative definition restricted to BRCA-specific codes yielded 65 carriers (1.1%), confirming the finding is not an artifact of code selection. The overwhelming majority of individuals carrying a high-risk variant therefore had no record of clinical genetic testing. Findings were robust to gene-panel choice. The ACMG SF v3.2 cancer subset yielded a higher apparent prevalence (2.68%), but more than half of carriers were heterozygous MUTYH (5,861 of 11,131); excluding MUTYH, prevalence was 1.29% (95% CI, 1.25–1.32) — close to the HBOC estimate (1.42%) — and documented testing among women carriers remained low (1.7% overall; 2.2% European-ancestry). Documented testing was ≈1.1–2.2% across all panel definitions, and the ancestry and socioeconomic patterns were unchanged (eTable 3).

### Disparities in Documented Testing by Ancestry and Socioeconomic Position

Among 250,071 women, a cancer diagnosis was the strongest correlate of documented testing (aOR, ∼9), consistent with diagnosis-triggered testing, and carrier status itself was not significantly associated with documented testing (aOR, 1.23; 95% CI, 0.92–1.65), underscoring that carriage rarely prompts testing in the absence of disease. Relative to European-ancestry women, African-ancestry women had markedly lower adjusted odds of documented testing in Model 1 (aOR, 0.32; 95% CI, 0.27–0.39), as did Admixed American women (aOR, 0.50; 95% CI, 0.43–0.58); East Asian, Middle Eastern, and South Asian estimates did not differ significantly (**Figure 2**; **eTable 2** in the Supplement).

**Figure 2.**
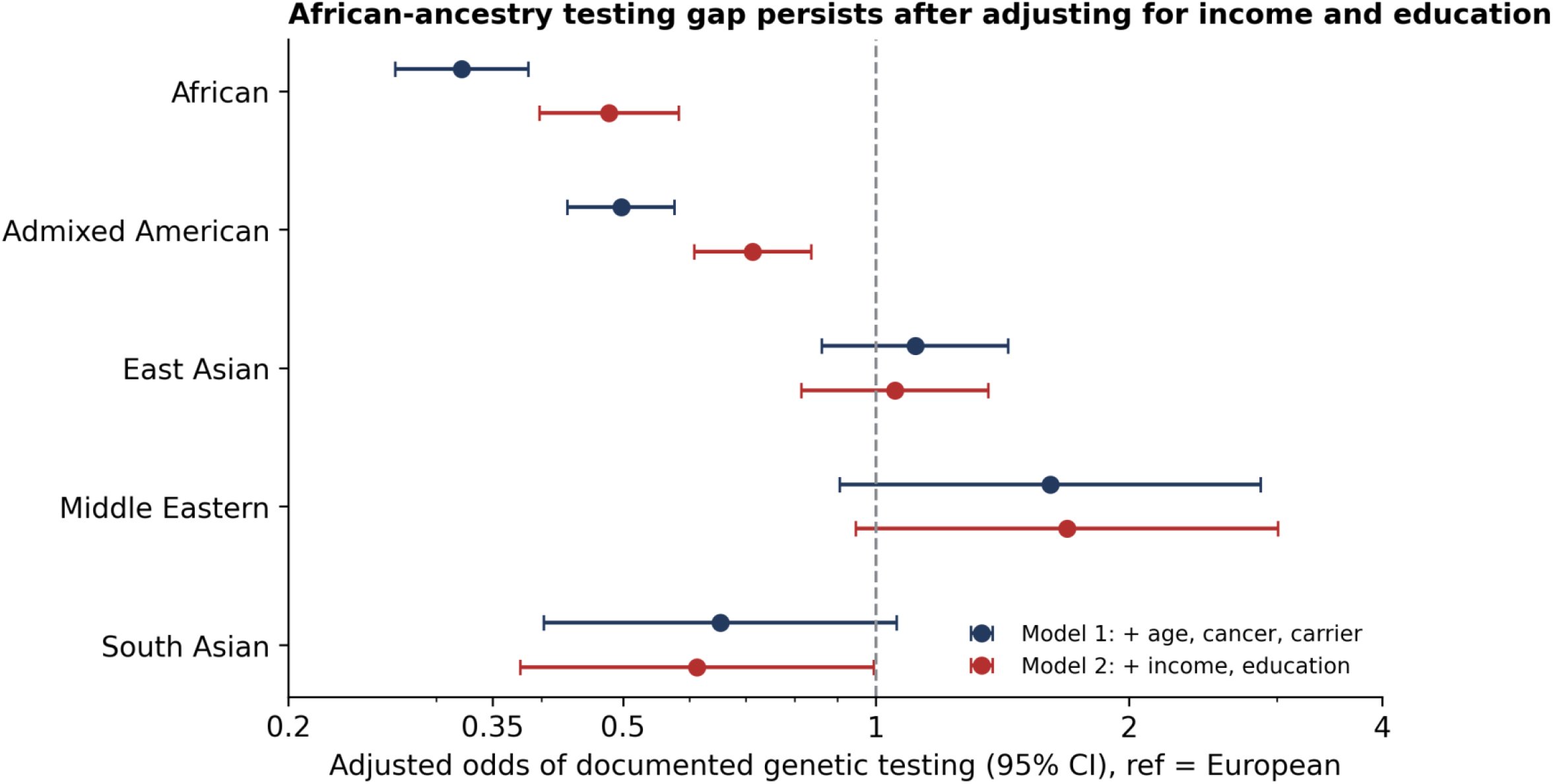
Adjusted odds of documented genetic testing by ancestry (reference, European). The African-ancestry gap attenuates but persists after adjustment for income and education.

After adjustment for income and education (Model 2), the African-ancestry gap attenuated but remained highly significant (aOR, 0.48; 95% CI, 0.40–0.58; P < .001), and the Admixed American difference, though further attenuated, also persisted (aOR, 0.71; 95% CI, 0.61–0.84; P < .001). Income and education were each independently and dose-dependently associated with documented testing (**Figure 3**): relative to household income >$100,000, odds declined monotonically to 0.46 (95% CI, 0.39–0.56) for income <$25,000; relative to a college degree, odds were 0.54 (95% CI, 0.45–0.66) for high-school education. Socioeconomic position thus partially attenuated, but did not fully explain, the ancestry differences in documented testing.

**Figure 3.**
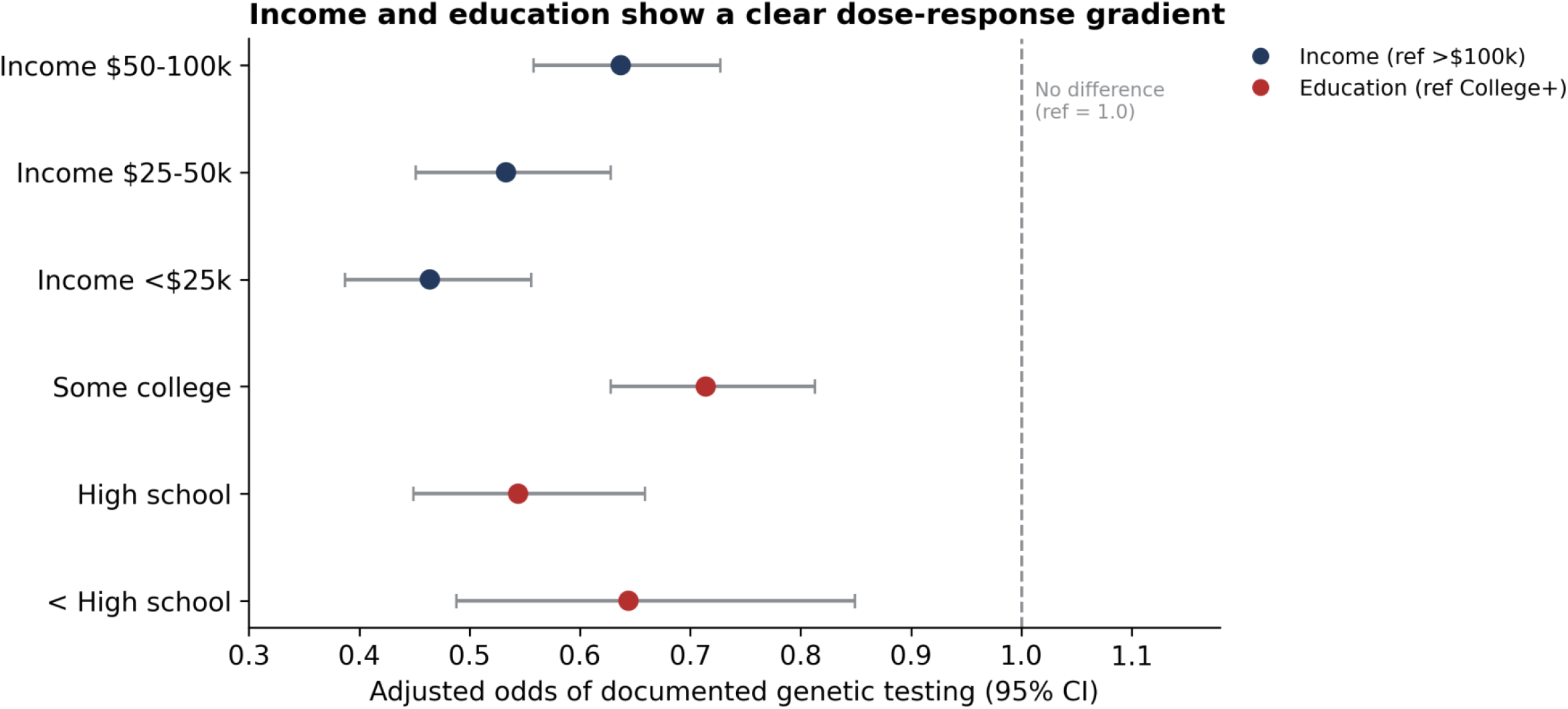
Adjusted odds of documented genetic testing by household income and educational attainment (Model 2), showing a dose-response gradient.

## Discussion

In a large, ancestrally diverse U.S. cohort, we found that P/LP HBOC variant carriers were present across all genetic ancestry groups at similar frequencies (∼1.4%), yet documented clinical genetic testing reached only a small minority of carriers. Among the disparities in documented testing, the African-ancestry deficit was the most striking and, importantly, persisted after accounting for income and education; the Admixed American gap was attenuated by socioeconomic position but also remained significant. These findings localize the carrier-identification shortfall and show that ancestry differences in documented testing are only partially explained by socioeconomic position.

Our prevalence estimates are concordant with prior All of Us analyses reporting broadly similar pathogenic-variant frequencies across ancestry groups[17], and with population-screening studies in which the great majority of BRCA carriers are never clinically identified[3 12]. Our contribution is to pair direct, sequencing-based carrier ascertainment with linked records of clinical testing in the same individuals, and to show that the residual African-ancestry gap cannot be reduced to socioeconomic differences. This pattern is consistent with literature implicating provider referral, awareness, and structural access—rather than patient resources alone—as central drivers of testing inequity[13-15], and it argues for interventions that do not depend on the existing referral pathway.

The dose-response associations of income and education with documented testing reinforce that socioeconomic position remains an independent barrier even where it does not fully explain ancestry differences. The strong association of a cancer diagnosis with testing, together with the only modest association of carrier status, indicates that testing in this cohort was largely reactive to disease rather than anticipatory of risk—precisely the pattern that population-based screening aims to change.

## Limitations

This study has several limitations. First, documented genetic testing was ascertained from EHR procedure codes, which incompletely capture testing performed outside the linked health systems; absolute testing rates are therefore underestimates, and the detection gap, while real and large, should be interpreted as documented testing. Differential EHR completeness across groups could bias disparity estimates in either direction. Second, carrier status reflected carriage of at least one alternate allele without zygosity resolution, and variant classification depended on ClinVar assertions, which evolve. Third, genetic ancestry is a coarse construct and does not capture the social and structural determinants it may proxy. Fourth, the cross-sectional design precludes temporal and causal inference. Finally, small numbers of tested carriers in non-European groups required suppression, limiting group-specific detection-rate estimates. The magnitude of under-ascertainment is substantial: in All of Us, structured genetic-testing codes appear in fewer than 1% of records even among patients with cancer[22], largely because most hereditary testing is performed by send-out reference laboratories whose results are not recorded as structured procedures. We therefore defined documented testing broadly (hereditary-cancer gene-analysis, multigene-panel [including proprietary laboratory analysis], and genetic-counseling codes); a conservative BRCA-specific definition (65 carriers; 1.1%) yielded the same conclusions. The gene-panel choice was also examined: our panel comprised the actionable NCCN HBOC genes, and a broader ACMG cancer panel raised apparent prevalence chiefly through non-actionable monoallelic MUTYH carriers and non– breast/ovarian syndromes; excluding MUTYH returned prevalence near the HBOC estimate, with documented testing rare throughout, indicating the detection gap is not an artifact of gene-panel selection. Notably, only 7 of the 29 ACMG cancer genes confer established breast and/or ovarian cancer risk (BRCA1, BRCA2, PALB2, TP53, PTEN, STK11, CDH1)[23]; the remainder define non-breast/ovarian syndromes, which, together with monoallelic MUYTH, accounted for most of the excess ACMG carrier count.

## Conclusions

High-risk HBOC variant carriers are distributed across all ancestry groups at similar frequencies, but documented clinical genetic testing identifies only a small fraction of them, and African-ancestry women face a testing gap that is not explained by socioeconomic position alone. Closing this gap will likely require population-level identification strategies that operate independently of current, referral-dependent pathways.

## Data Availability

All data produced are available online at All of Us Research Program.

## Article Information

## Acknowledgments

We gratefully acknowledge All of Us participants for their contributions, without whom this research would not have been possible, and the National Institutes of Health’s All of Us Research Program for making the participant data available.

## Author Contributions

S.Y.S had full access to all the data in the study and takes responsibility for the integrity of the data and the accuracy of the data analysis. Concept and design: S.Y.S. Acquisition, analysis, or interpretation of data: S.Y.S Drafting of the manuscript: S.Y.S and S.H. Critical revision of the manuscript for important intellectual content: S.Y.S. Statistical analysis: S.Y.S and S.H. Supervision: S.Y.S.

## Data Availability Statement

The data underlying this study are available to authorized researchers through the All of Us Research Workbench (Controlled Tier; Curated Data Repository v8/C2024Q3R9) at https://www.researchallofus.org. Access requires registration, an institutional Data Use and Registration Agreement, and required training. The authors cannot redistribute individual-level data; all results are reported in aggregate with small-cell suppression (counts <20) per the All of Us Data and Statistics

## Dissemination Policy

Analysis code is available from the corresponding author on reasonable request.

## Ethics Approval and Consent

This study used de-identified data from the All of Us Research Program under the program’s approved governance and institutional review board oversight. All All of Us participants provided informed consent to the program.

## Reporting Guideline

This cross-sectional study is reported in accordance with the STROBE (Strengthening the Reporting of Observational Studies in Epidemiology) guideline; the completed checklist is provided as supplementary material.

